# Ambient Air Pollution and Smell Function: Evidence from the National Geographic Smell Survey

**DOI:** 10.1101/2025.06.10.25329361

**Authors:** Vicente Ramirez, Sandie Ha, Valentina Parma, Pamela Dalton, Joel D. Mainland, Danielle R. Reed

## Abstract

**Background:** Olfactory sensory neurons are located outside the brain, allowing them to detect environmental chemicals. However, this comes at the cost of exposure to potential toxins, which may decrease olfactory function.

**Methods:** We sought to assess the association between olfactory function and air pollution, measured by the National Geographic Smell Survey and data from the Environmental Protection Agency. We examined the effects of air pollution exposure on perceived odor intensity and identification ability across 97,087 survey respondents assigned air pollution exposures at the zip code level.

**Results:** The results show that NO_2_, SO_2_, and O_3_ were associated with slight alterations in olfactory acuity, and these relationships differed by odor and the type of pollutant. Specifically, increases in annual mean concentration by a standard deviation of NO_2_ (OR=0.97; CI=0.96-0.97) and SO_2_ (OR=0.99; CI=0.98-1.00) were associated with reduced odor identification ability. Conversely, increases in O_3_ concentration were associated with a slight increase in olfactory ability (OR=1.01; CI=1.00-1.02).

**Conclusions:** Air quality is associated with olfactory health, underscoring the need to investigate the mechanisms driving pollution-induced impairment.

## Introduction

Environmental pollution has become one of the most pressing issues in global health, as growing evidence links ambient environmental exposures to multiple health outcomes, including cardiovascular, respiratory, mental health, and neurodegenerative disorders^1–3^.

The olfactory epithelium is positioned at the interface of the internal and external milieu, and its unique, direct connection between olfactory sensory neurons and the brain renders it especially susceptible to harmful environmental exposures. As demonstrated in animal models, air pollutants can reach the brain through this olfactory route, disrupting neural function and promoting neurodegeneration^4,5^. In humans, olfactory dysfunction is an early marker for Alzheimer’s and Parkinson’s diseases, cognitive decline, longevity, and overall well-being—all of which have demonstrated associations with air pollution^6–11^. It is plausible that toxic environmental exposures may compound declines in olfactory function, indicating early health deterioration.

Early studies on the relationship between ambient air pollution and olfactory function primarily compared populations from high- and low-pollution areas. Two reports compared olfactory function in adults exposed to high pollution levels in Mexico City to adults in the less polluted Mexican State of Tlaxcala^12,13^. Using ecological [commercial beverage^13^] and standardized stimuli^12^, these studies found lower odor thresholds, higher odor discrimination scores, and mild microsmia in 12% of residents, compared to 35.5% of Mexico City residents with mild to severe microsmia^12,13^. Calderón-Garcidueñas et al. performed autopsies on 35 Mexico City residents and nine controls (including children), reporting structural abnormalities in olfactory bulbs, the presence of ultrafine particulate matter in olfactory bulbs, and of amyloid-β 42 and α-synuclein in the olfactory nerves and bulbs of the cadavers from Mexico City but not in controls^12^. These findings suggest that olfactory deficits and neurodegenerative biomarkers emerge early during development and may be influenced by environmental pollutant exposure.

Even though an increasing body of research links exposure to pollutants (e.g., airborne particulates, gaseous phase pollutants, heavy metals) with olfactory dysfunction^6,11,14–17^, the relationship between air pollution, olfactory and general health remains poorly understood, as it is often conducted in studies with small sample size.

The landmark National Geographic Smell Survey (NGSS), distributed in the September 1986 issue of the National Geographic Magazine, offers a psychophysical, multifunction assessment of olfactory ability, measuring odor detection, intensity, and identification, measures that reflect peripheral (nose) and central (brain) olfactory function^18,19^. With data from over 1.2 million individuals in the United States (U.S.), the NGSS is one of the largest databases with psychophysical measures of human olfactory function^20^ .It includes geographic indicators (zip codes) so we can examine the relationship between ambient air pollution and olfactory health when combined with U.S. Environmental Protection Agency (EPA) Air Quality Surveillance data^21^. The late 1980s were a time of heightened air pollution, just before the implementation of key regulatory measures such as the 1987 Montreal Protocol and the 1990 Clean Air Act Amendments^22,23^.

Our study aims to explore the impact of common specific air pollutants in the U.S. on odor intensity and identification, contributing to the growing body of literature on environmental influences on olfactory function. We hypothesize that higher average annual pollution exposure is associated with decreased olfactory performance.

## Methods

### Sampling Strategy

The NGSS was mailed to every *National Geographic* magazine subscriber. **Figure S1** shows the exclusion criteria applied to this convenience sampling, which resulted in a final analytic sample of 97,087 U.S. individuals with complete olfactory and air pollutant exposure data.

### Olfactory Assessment

The NGSS, a self-administered "scratch-and-sniff" booklet, employed a panel of 6 odors (**Table S1**)^20^ to obtain multiple dimensions of olfactory function, including odor intensity and identification. Intensity was rated on an ordinal scale from 1 (weak) to 5 (strong). Odor identification was assessed using a 12-choice panel of 10 odor descriptors, a “no odor” option, and an “other” option. Identification accuracy was coded as a binary variable, indicating whether the participant correctly matched each odor. A non-response was considered incorrect if a participant did not respond to an odor, but responded to other odors. Otherwise, individuals who did not respond to all odors were removed from the study sample. A total number of correct identifications was scored on a 0-6 scale, summing correct responses across six trials. **See Figure S2 for the psychophysical odor portion of the NGSS.**

### Air Pollution Measures

The EPA has standardized air quality data collections since 1980. To determine exposure to five criteria pollutants (PM_10_, Pb, SO_2_, NO_2_, and O_3_), we used the 1986 annual concentration data from U.S. air monitoring sites^24^

### Linking Pollutant Exposure to Olfactory Performance

The NGSS provides participant residence based on zip code, while the EPA monitors air quality at specific locations with latitude and longitude coordinates. To link these datasets, we used shapefiles for Zip Code Tabulation Areas (ZCTAs) from the 2000 U.S. Census (which are the earliest ZCTAs available) to extract geographic coordinates for zip code boundaries and centroids across the U.S^25^. We projected these geographical data using the R packages *rgdal*, *rgeo*, and *sf,* and a 30 km spatial buffer was created around zip code centroids^26–28^. Monitoring sites within 30km of a centroid were assigned to that zip code.

Zip codes with concentrations for all five pollutants of interest were utilized in our analysis. Crosswalk files were used to fix changes in zip code assignment and boundaries between 1990 and 2000, and merged with the NGSS zip code data. This updated NGSS was then merged with the ZCTA geocoded monitoring sites data, and annual concentrations of air pollutants for each participant were assigned based on their reported zip code. Participants missing data in any of the variables of interest were removed (**Figure S1**).

The final sample included 97,087 individuals, each assigned annual-averaged concentrations for PM_10_, Pb, SO_2_, NO_2_, and O_3_. Table 1 details descriptive statistics on demographic variables and average assigned air pollution exposures.

**Table 1.** Description of demographic features of the NGSS sample analyzed.

|  | **n = 97087** |
| --- | --- |
| **Age (continuous)** | 41.7 (15.8) |

| **Mean (SD)** |  |  |
| --- | --- | --- |
| **Age (stratified)**<br>**N (%)** | 10-20 | 6999 (7.2%) |
|  | 20-35 | 31527 (32.5%) |
|  | 36-50 | 30084 (31%) |
|  | 51-65 | 20352 (21%) |
|  | 66-80 | 7621 (7.8%) |
|  | 80+ | 504 (0.5%) |
| **Sex**<br>**N (%)** | Female | 39211 (40.4%) |
|  | Male | 57876 (59.6%) |
| **Race/Ethnicity**<br>**N (%)** | White | 90631 (93.4%) |
|  | Black | 1306 (1.3%) |
|  | Hispanic | 1147 (1.2%) |
|  | Asian | 1118 (1.2%) |
|  | American Native | 140 (0.1%) |
|  | Other | 374 (0.4%) |
|  | No Response | 2371 (2.4%) |
| **Smoke Status**<br>**N(%)** | Non-smoker | 78355 (80.7%) |
|  | Smoker | 18732 (19.3%) |
| **Self Rated Smell (1-5)**<br>**N(%)** | 1 | 623 (0.6%) |
|  | 2 | 2502 (2.6%) |
|  | 3 | 20192 (20.8%) |
|  | 4 | 41468 (42.7%) |
|  | 5 | 31435 (32.4%) |
|  | No Response | 867 (0.9%) |
| **Average Annual**<br>**Pollution Exposure**<br>**Mean (SD)** | PM<sub>10</sub> µg/m<sup>3</sup> | 39.9 (6.4) |
|  | Pb µg/m<sup>3</sup> | 0.2 (0.1) |
|  | O<sub>3</sub> ppm | 0.05 (0.007) |
|  | NO<sub>2</sub> ppb | 27.8 (8.1) |
|  | SO<sub>2</sub> ppb | 9.8 (3.9) |

**Note:** PM<sub>10</sub>, particulate matter with aerodynamic diameter ≤10 micrometers; Pb, lead; O<sub>3</sub>, ozone; NO<sub>2</sub>, nitrogen dioxide; SO<sub>2</sub>, sulfur dioxide; ppm, parts per million; ppb, parts per billion; µg/m<sup>3</sup>, micrograms per cubic meter; N, number of individuals from the sample; SD, standard deviation.

### Statistical Analysis

Multiple single-pollutant regression models were utilized to assess the association between each pollutant, the perceived intensity of each odor, and the correct identification of odors. All models included mean annual pollutant exposure as an independent variable and covariates [i.e., sex, age, and smoking status, known modulators of olfactory function^20,29,30^]. Mean annual pollutant exposures were z-scored, and age was centered around its mean. We utilized partial proportional odds logistic regression models to assess the association between intensity ratings and air pollution exposure. Models were fit in STATA 16 using the GOLOGIT2 library and the -autofit option, which optimizes constraints on explanatory variables to meet the proportional odds assumption^31,32^. Multilevel logistic regression models examined the relationship between participants’ exposure to individual air pollutants and identification scores, with random-intercepts grouped to each individual across the 6-odor identification trials. Similarly, logistic regression models were used to assess the identification of individual odors. We applied the Benjamini-Hochberg correction to account for the False Discovery Rate on multiple comparisons^33^. All analyses were run in STATA 16, R 4.3, and RStudio 2024.12.1. Figures were created using *R* and the *ggplot2* package.

## Results

### Weak and Inconsistent Associations Between Air Pollution and Odor Intensity

The association between air pollution and perceived odor intensity is modest and variable in direction and magnitude. **Figure 1** shows single-pollutant associations with odor intensity by outcome threshold (how intense the odor was rated from 1 to 5, **see Supplementary Files** for model, covariate-adjusted estimates). A standard deviation (SD) increase in PM_10_ increased the odds of intensity ratings >1 and >4 androstenone.

**Figure 1.**
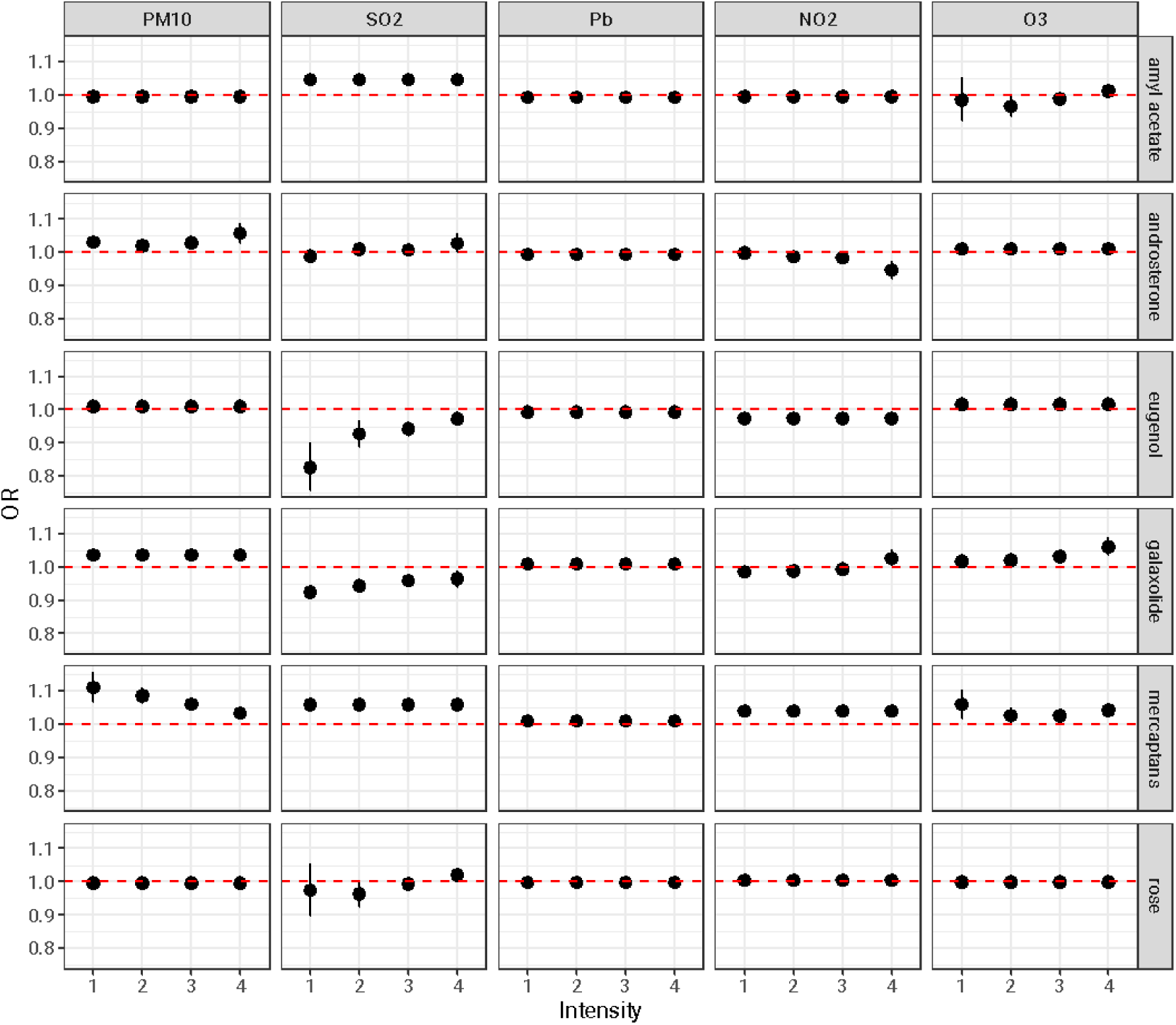
Generalized Ordinal Logistic Regression (Partial Proportional Odds Model) of Intensity Ratings. Perceived intensity rating cut points are listed on the X-axis, and odds ratios on the Y-axis. The odds ratio for each intensity cutpoint is the odds of rating greater than that cutpoint (e.g., Rating >1 vs 1, Rating >2 vs 1 or 2). Black bars represent the 95% confidence interval. The red line represents the null assumption, and represents the baseline odds ratio (e.g., OR=1)

Conversely, the effect of NO_2_ was not significant for rating androstenone intensity above 1, 2, or 3 but showed a modest effect on higher intensity rating thresholds with decreased odds of rating >4.

A one SD increase in exposure to PM_10_ was associated with higher intensity ratings of galaxolide across all rating thresholds. Similarly, an association between O_3_ exposure and galaxolide ratings was found, but this effect was only significant for rating thresholds >3 and >4. A one SD increase in SO_2_ concentration decreased the odds of higher-intensity ratings for galaxolide by ∼3-8% across rating thresholds between 1 and 3. The relationship diminished after adjusting for multiple testing at ratings threshold >4.

The strongest associations were found between SO_2_ and NO_2_ with eugenol intensity, though the effects varied across thresholds. An SD increase in SO_2_ concentration led to a 17.5% decrease in the odds of rating the intensity >1 and a 3% decrease in rating the intensity >4. An SD increase in NO_2_ reduced the odds of higher intensity eugenol ratings by ∼3% across all rating thresholds.

The relationship between O_3_ and amyl acetate was mixed: we found a negative relationship was for ratings >2 and positive for ratings >4, however, the significance of the relationship at rating threshold >2 diminished after adjusting p-values for multiple testing. An SD increase in SO_2_ concentration was associated with a 4.7% increase in the odds of rating amyl acetate intensity higher across all thresholds. All pollutants, except Pb, were associated with higher intensity ratings of mercaptans per SD increase in concentration. No pollutant had significant associations with intensity ratings of rose.

### Strong Association between Pollutant Exposure and Overall Odor Identification Ability

The results for odor identification are modest and show a mixed relationship between air pollution exposure and odor identification ability (**Figure 2**).

**Figure 2.**
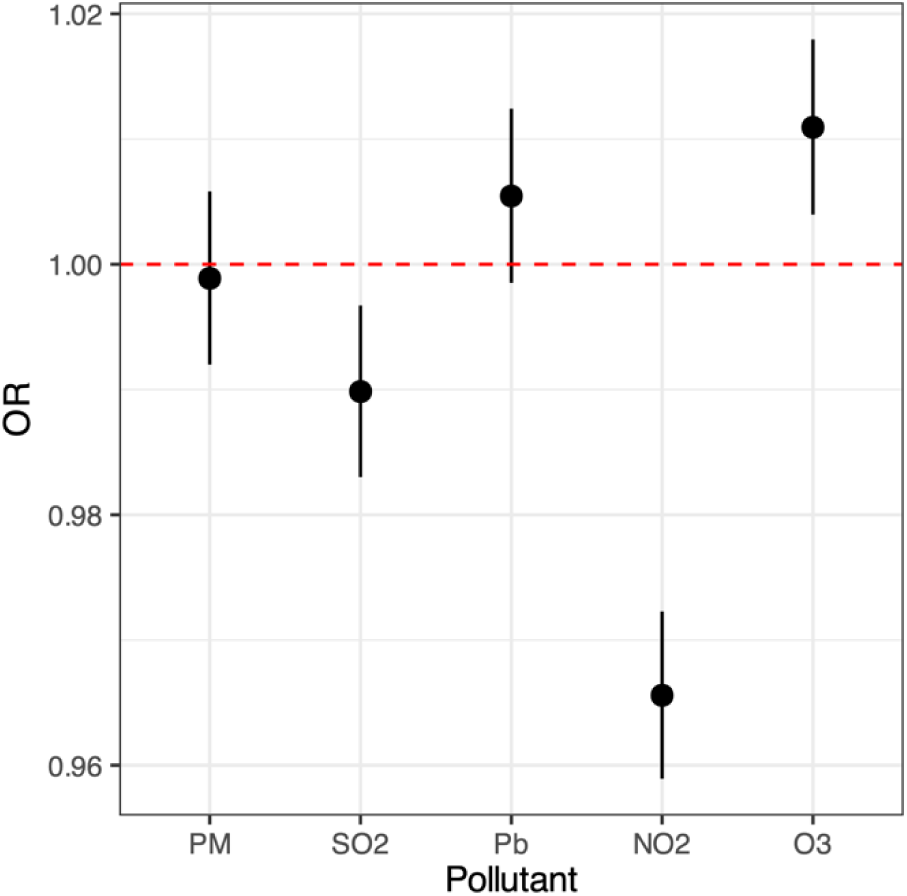
Binomial Logistic Regression for Correct Identification of Odors. The odds ratios for correctly identifying an odor over the six odor trials are plotted for each pollutant. Black bars represent the 95% confidence interval. The red line represents the null assumption and the baseline odds ratio.

Decreased odor identification performance was associated with an SD increase in NO_2_ (OR=0.97; CI=0.96-0.97) and SO_2_ (OR=0.99; CI=0.98-1.00) concentrations. However, an SD increase in O_3_ concentration was associated with a slight increase in olfactory ability (OR=1.01; CI=1.00-1.02). No associations were found between PM_10_ and Pb with overall odor identification ability.

After removing galaxolide and androstenone, for which specific anosmias have been documented^34^ the associations between olfactory identification ability and pollutant exposures remained for NO_2_ and SO_2_, but not for O_3_. Additionally, a small positive association was found between olfactory ability and Pb (OR=1.01; CI=1.00-1.02).

A binary logistic regression for identifying individual odors reveals heterogeneity in the relationship between pollution exposures and olfactory identification of individual odors. Among the strongest relationships, a moderate association between NO2 and SO2 and eugenol identification revealed a 12.1% and 10.3% decrease in the odds of correctly identifying eugenol per SD increase of NO_2_ and SO_2,_ respectively. Statistically significant associations were observed in both negative and positive directions across all odors and pollutants, as documented in **Figure 3**.

**Figure 3.**
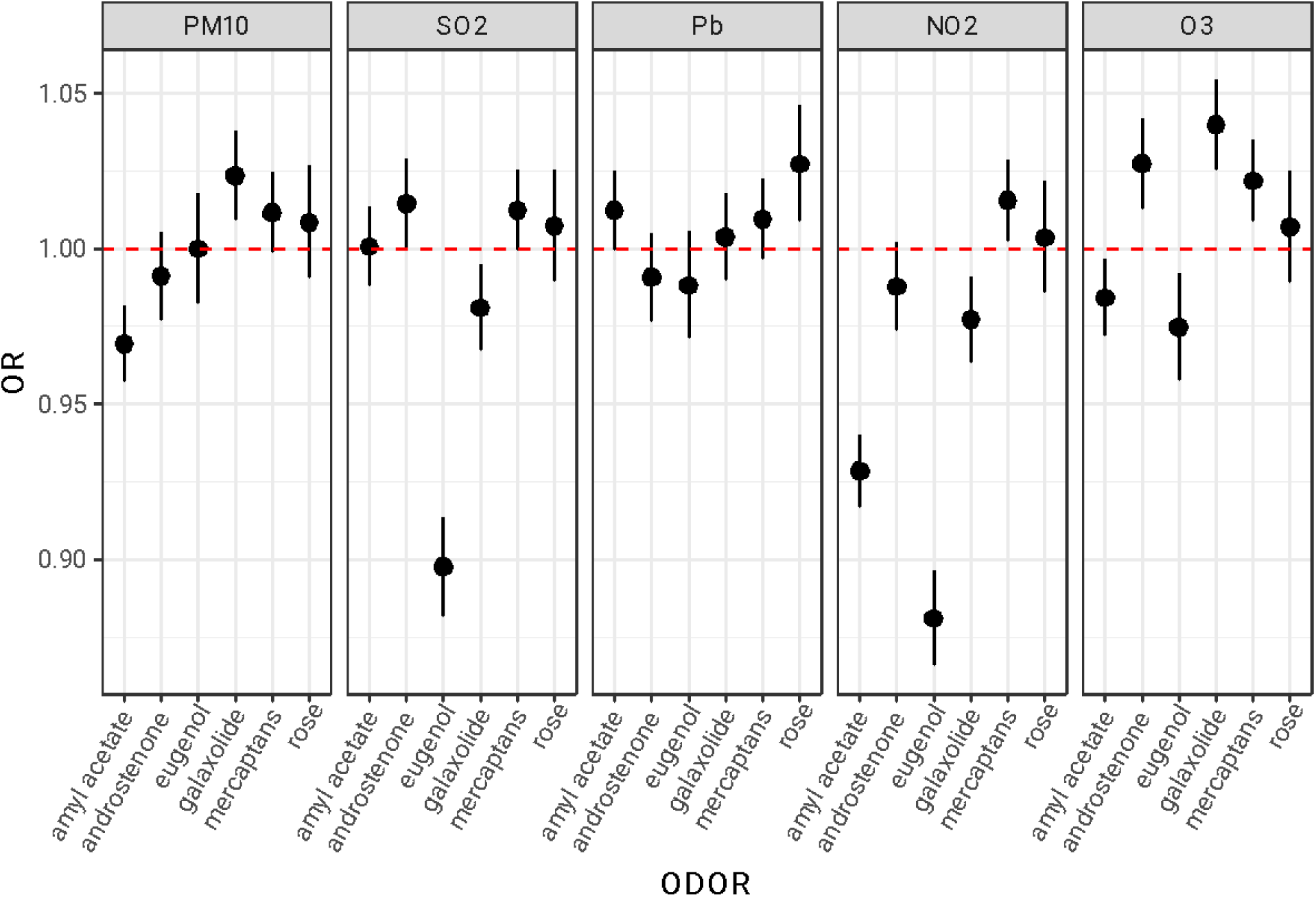
Logistic Regression for Correct Identification By Odor. **Note.** Odds ratios for correctly identifying individual odors vs the pollutant. Black bars represent the 95% confidence interval. The red line represents the null assumption, and represents the baseline odds ratio (e.g., OR=1)

## Discussion

### Main Findings

We assessed the association between air pollution exposure and olfactory function in a large, population-based sample of 97,087 U.S participants. We utilized a psychophysical smell test to evaluate multiple dimensions of olfactory function. Our findings were mixed: exposure to certain pollutants, such as NO_2_ and SO_2_, was associated with a significant decline in olfactory performance, whereas other pollutants, such as Pb, demonstrated no measurable impact at the observed exposure levels. While the evidence supports a relationship between air pollution and olfactory function, the observed effects were modest overall. The strongest pollutant-specific associations were limited to single odors or specific intensity thresholds.

Overall, NO_2_ showed the most consistent and strongest estimates for its negative effect on olfactory function, followed by SO_2_. These results align with prior literature linking NO_2_ to adverse effects on physiological processes and negative health outcomes^6,35,36^.

However, some findings diverged from our hypotheses, suggesting potential protective effects of certain pollutants or interactions between pollutants. For example, O_3_ exposure was associated with increased intensity ratings for multiple odorants but concurrently linked to reduced identification of specific odorants (e.g., eugenol and amyl acetate).

Accepting these findings as valid suggests that the effects of air pollution on olfactory function are heterogeneous, likely influencing distinct olfactory pathways via diverse mechanisms. However, the current data do not allow us to elucidate these mechanisms fully, but suggest that reduced odor identification may provide indirect evidence that central olfactory pathways are vulnerable to gaseous pollutants. Air pollutants can adversely affect the olfactory system through multiple direct and indirect mechanisms: i) exposure damaging the olfactory epithelium, causing structural changes and impairing function^4,15,37^ ; ii) pollutant-triggered neuroinflammation and oxidative stressor^8,15^; iii) translocation from nasal mucosa to central nervous system, bypassing the blood-brain barrier^4,9^; iv) pollutant-triggered mitochondrial dysfunction, which compromises homeostasis^38^. Future research is needed to clarify how gaseous pollutants affect smell function.

To incorporate the findings that exposure to some pollutants, like O_3_, may be a protective factor for olfactory ability, we note that air pollution effects are contextual and interdependent. For example, the inverse concentration relationship between O_3_ and NO_2_ can confound observed health effects, as these relationships are highly dependent on local environmental conditions and exposure contexts and can influence the spatial variability of these pollutants^39–41^. For example, the NGSS survey was disseminated in September 1986, which aligns with the US cold season, when NO_2_ levels are typically high and O_3_ levels are low. Notably, O_3_ at low concentrations has been associated with protective effects on health^42^.

While most of these pollutants’ effects are modest, the patterns indicate that alterations in olfactory function may be partially attributable to environmental exposures, warranting further investigation into pollutant-specific and context-dependent effects.

### What is known

Recent studies have suggested that environmental exposures alter olfactory function. Evidence has accumulated linking olfactory dysfunction to exposure to various pollutants such as airborne particulates, gaseous phase pollutants, and heavy metals^6,14,15^. Our analysis found that gaseous pollutants produced the largest effects, with NO_2_ revealing the most consistent findings. This result agrees with previous studies that have associated urban air pollutants such as NO_2_ with olfactory dysfunction, particularly in elderly individuals^6,43^. Zhang et al. found an association between anosmia and long-term exposure to PM_2.5_ by examining the ICD9/10 diagnostic codes^44^. Scussiatto et al. demonstrated that ambient exposure to PM_10_ was associated with decreased smell identification scores^17^ We did not find such associations; instead, we found that the PM_10_ was often protective or irrelevant. While unexpected, this is consistent with recent findings from Oleszkiewicz et al., which suggested that airborne particulates were associated with increased sensitivity^16^.

Our findings align with previous work, which associated NO_2_ exposure with poor olfactory function in older urban-dwelling adults^6^. With this in mind, we conclude that increased exposure to NO_2_ and SO_2_, such as simply living near a busy roadway, may lead to increased risks of a decline in olfactory function^8,15,45^.

### What this study adds

To date, this dataset represents the largest multi-dimensional olfactory database matched with environmental data, providing a unique opportunity and, critically, a model for future exploration of inter-individual variability in olfactory function and pollutant exposures.

Our findings reveal that air pollution is more consistently associated with alterations in olfactory identification, expected to be mediated by central mechanisms, rather than olfactory intensity ratings, a proxy for olfactory sensitivity, linked to function in the peripheral olfactory system.

This study generates new hypotheses regarding the effects of specific air pollutants on olfactory function. It underscores the need for high-resolution population-based assessments of multiple olfactory dimensions to be conducted longitudinally.

### Limitations

A few limitations warrant caution in interpreting the results. First, the observed differences in effects between odor identification and intensity may be attributed to methodological factors. The 1-5 scale used for intensity ratings reduces granularity (i.e., each step representing a 20% change), likely obscuring subtler effects and introducing variability due to inter-rater subjectivity. Additionally, the scratch-and-sniff method for odor delivery lacks standardization: the pressure applied during scratching and the distance during sniffing might contribute to variability in intensity ratings. However, because odor concentrations exceeded typical detection thresholds, these factors likely have a smaller, though non-negligible, impact on odor identification.

Second, although this sample size allows for powerful observations, using zip codes to assign air pollution exposure introduces bias as it does not account for spatial variability when assigning exposures. Annual averages in air pollution concentration could have masked shorter periods of peak exposures that may have heightened risks. Selection bias is also possible, as we only included people living in zip codes with air monitors within 30 km. Lastly, the study sample may not fully represent the general population because it was self-selected.

## Conclusions

Our findings indicate that significant effects on human olfaction were observed even at pollutant concentrations below the U.S. National Standards, which have implications for human health^46^.

## Data Availability

All data used in this study are publicly available.
Zip code tabulation files are publicly available through the US Census Bureau FTP
repository at https://www2.census.gov/geo/tiger/TIGER2010/ZCTA5/2000/.
Air monitoring sites and pollution data are publicly available through the US EPA Air
Quality Service at https://aqs.epa.gov/aqsweb/airdata/download_files.html
The National Geographic Smell Survey data is publicly available via Kaggle.
https://www.kaggle.com/datasets/jmainland/national-geographic-smell-survey

https://www.kaggle.com/datasets/jmainland/national-geographic-smell-survey

## Data Availability

All data used in this study are publicly available.

Zip code tabulation files are publicly available through the US Census Bureau FTP repository at https://www2.census.gov/geo/tiger/TIGER2010/ZCTA5/2000/.

Air monitoring sites and pollution data are publicly available through the US EPA Air Quality Service at https://aqs.epa.gov/aqsweb/airdata/download_files.html

The National Geographic Smell Survey data is publicly available via Kaggle. https://www.kaggle.com/datasets/jmainland/national-geographic-smell-survey

---

**Figure S1.**
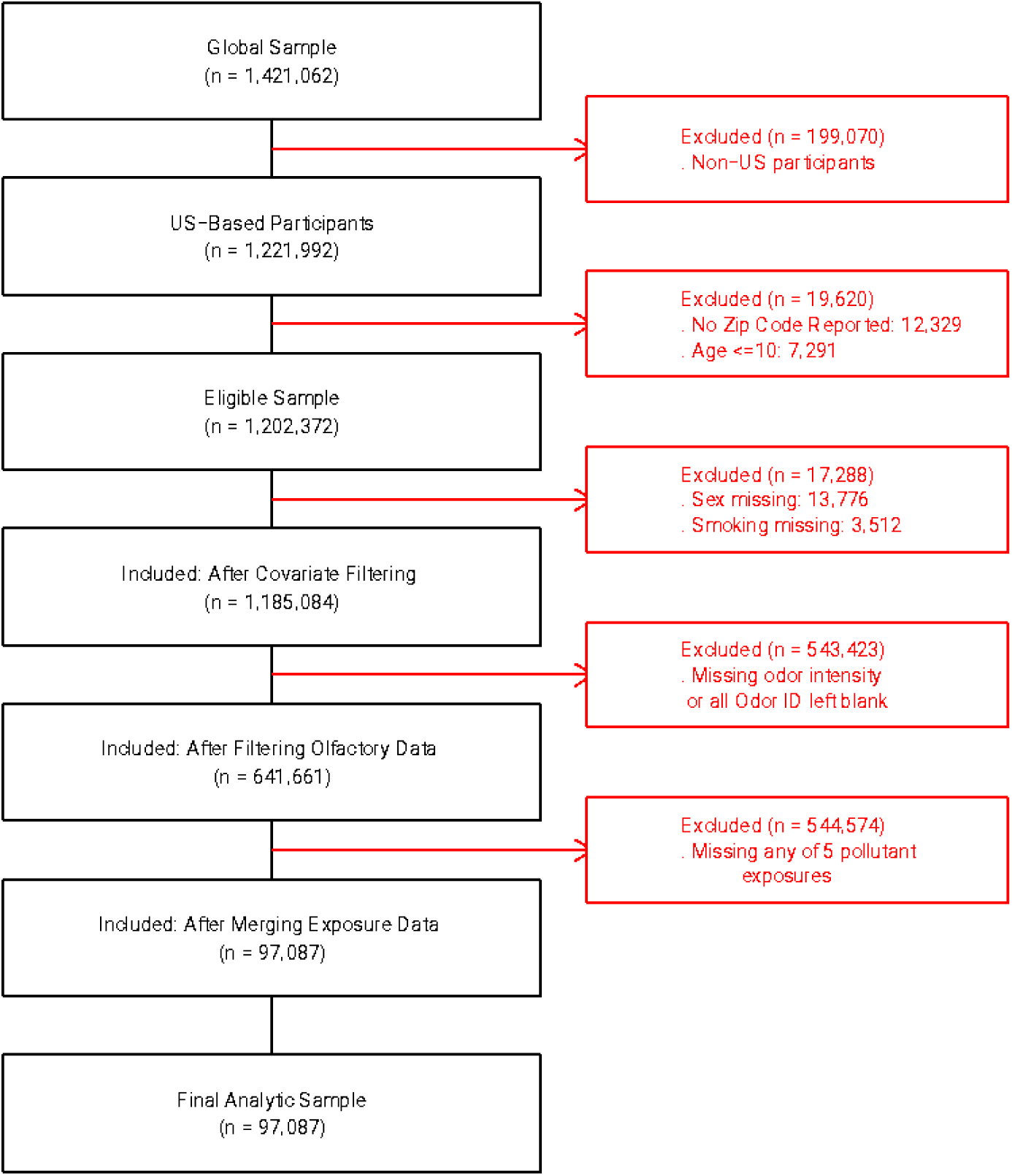
Flow diagram depicting the progression of participants through a study. **Note:** reproduced based on STROBE guidelines for reporting observational studies (von Elm et al., 2007).

**Figure S2.**
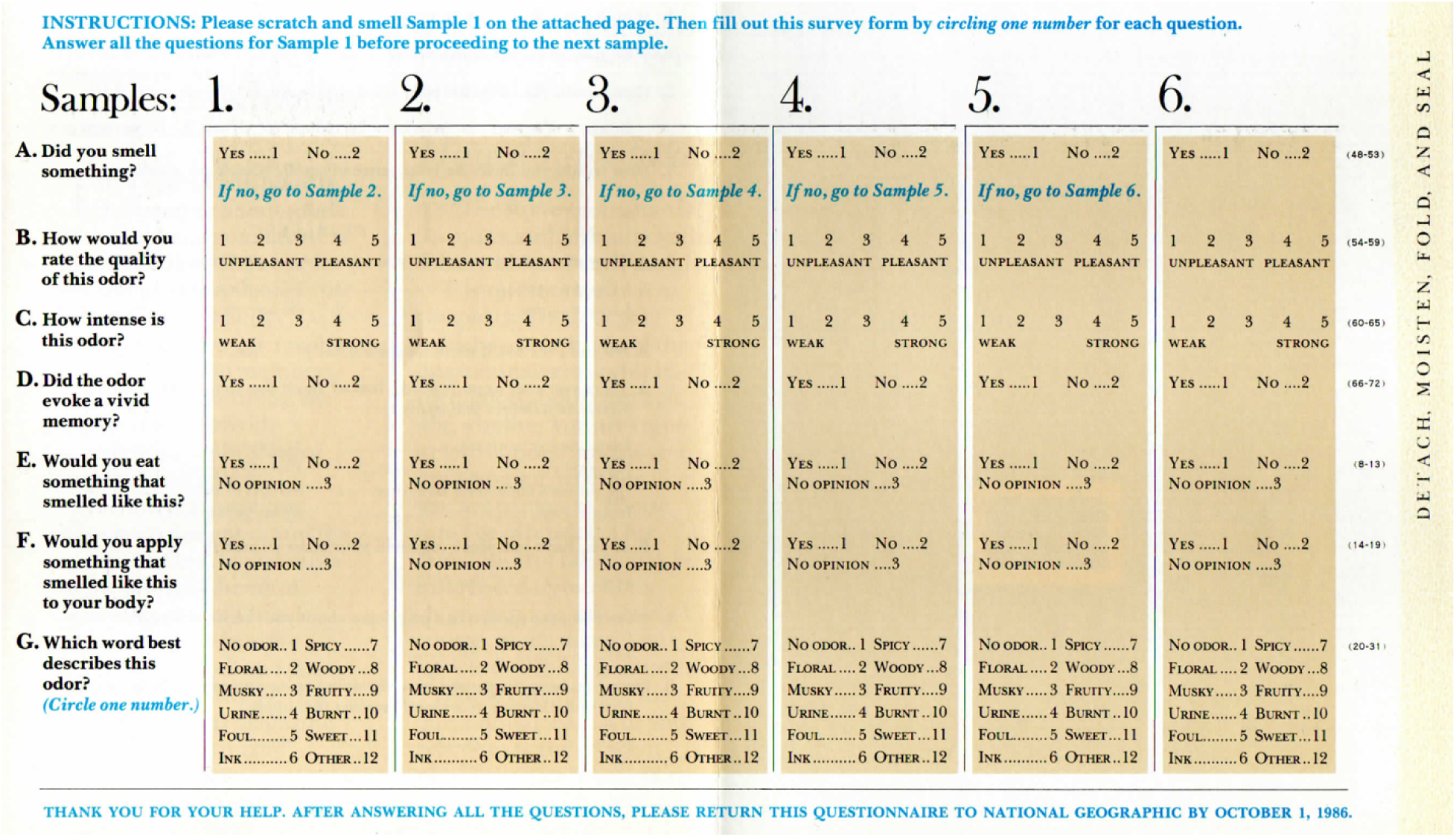
The Psychophysical Assessment in the National Geographic Smell Survey. Odor intensity and identification are assessed in portions C (odor intensity) and G (odor identification) of the NGSS.

**Table S1.**
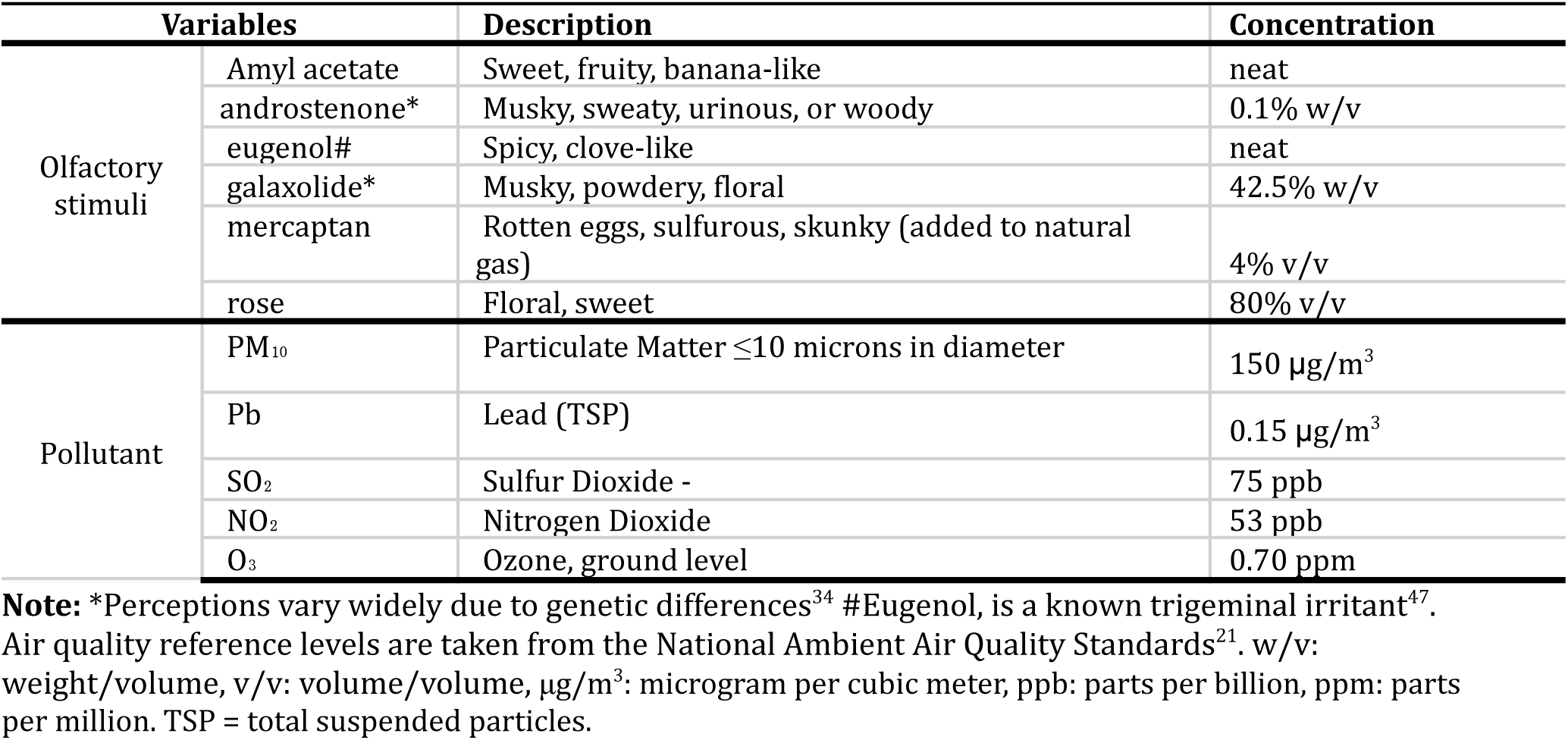
Odor stimuli in the NGSS and National Ambient Air Quality Standards for the Pollutants Measured.

**Alt-text Figure 1**: A grid of small multiple plots showing odds ratios (OR) with 95% confidence intervals across odor intensity cut point thresholds (>1 to >4) for six odors (amyl acetate, androsterone, eugenol, galaxolide, mercaptans, and rose) and five air pollutants (PM_10_, SO₂, Pb, NO₂, and O₃). Each panel represents the association between a specific odor and pollutant across increasing intensity levels. The y-axis represents the odds ratio (OR), and the x-axis represents odor intensity (1 to 4). Black dots with error bars represent the odds ratio (OR) estimates and confidence intervals. A red dashed horizontal line at OR = 1.0 indicates the null effect.

**Alt-text Figure 2:** A plot showing the odds ratio (OR) with 95% confidence intervals across five air pollutants (PM_10_, SO₂, Pb, NO₂, and O_3_ ). The y-axis represents the odds ratio (OR), and the x-axis represents odor intensity (1 to 4). Black dots with error bars represent the OR estimates and confidence intervals. A red dashed horizontal line at OR = 1.0 indicates the null effect.

**Alt-text Figure 3**: A panel of plots showing the odds ratio (OR) with 95% confidence intervals across five air pollutants (PM_10_, SO₂, Pb, NO₂, and O_3_). The y-axis represents the odds ratio (OR), and the x-axis represents odor tested (amyl acetate, androsterone, eugenol, galaxolide, mercaptans, and rose). Black dots with error bars represent the odds ratio (OR) estimates and confidence intervals. A red dashed horizontal line at OR = 1.0 indicates the null effect.

## References

1. de Bont J, Jaganathan S, Dahlquist M, Persson Å, Stafoggia M, Ljungman P. Ambient air pollution and cardiovascular diseases: An umbrella review of systematic reviews and meta-analyses. Journal of Internal Medicine. 2022;291(6):779–800. doi:10.1111/joim.13467

2. Kim H, Kim WH, Kim YY, Park HY. Air Pollution and Central Nervous System Disease: A Review of the Impact of Fine Particulate Matter on Neurological Disorders. Front Public Health. 2020;8:575330. doi:10.3389/fpubh.2020.575330

3. Losacco C, Perillo A. Particulate matter air pollution and respiratory impact on humans and animals. Environ Sci Pollut Res. 2018;25(34):33901–33910. doi:10.1007/s11356-018-3344-9

4. Calderon-Garciduenas L, Maronpot RR, Torres-Jardon R, et al. DNA Damage in Nasal and Brain Tissues of Canines Exposed to Air Pollutants Is Associated with Evidence of Chronic Brain Inflammation and Neurodegeneration. Toxicol Pathol. 2003;31(5):524–538. doi:10.1080/01926230390226645

5. Oberdörster G, Sharp Z, Atudorei V, et al. Translocation of inhaled ultrafine particles to the brain. In: Inhalation Toxicology. ; 2004. doi:10.1080/08958370490439597

6. Adams DR, Ajmani GS, Pun VC, et al. Nitrogen dioxide pollution exposure is associated with olfactory dysfunction in older U.S. adults: Nitrogen dioxide and olfactory dysfunction. Int Forum Allergy Rhinol. 2016;6(12):1245–1252. doi:10.1002/alr.21829

7. Calderón-Garcidueñas L, Reed W, Maronpot RR, et al. Brain Inflammation and Alzheimer’s-Like Pathology in Individuals Exposed to Severe Air Pollution. Toxicologic Pathology. 2004;32(6):650–658. doi:10.1080/01926230490520232

8. Calderón-Garcidueñas L, Solt AC, Henríquez-Roldán C, et al. Long-term Air Pollution Exposure Is Associated with Neuroinflammation, an Altered Innate Immune Response, Disruption of the Blood-Brain Barrier, Ultrafine Particulate Deposition, and Accumulation of Amyloid β-42 and α-Synuclein in Children and Young Adult. Toxicologic Pathology. 2008;36(2):289-310. doi:10.1177/0192623307313011

9. Garcia GJM, Schroeter JD, Kimbell JS. Olfactory deposition of inhaled nanoparticles in humans. Inhalation Toxicology. Published online 2015. doi:10.3109/08958378.2015.1066904

10. Grashow R, Sparrow D, Hu H, Weisskopf MG. Cumulative lead exposure is associated with reduced olfactory recognition performance in elderly men: The Normative Aging Study. NeuroToxicology. 2015;49:158–164. doi:10.1016/j.neuro.2015.06.006

11. Lucchini RG, Guazzetti S, Zoni S, et al. Neurofunctional dopaminergic impairment in elderly after lifetime exposure to manganese. Neurotoxicology. 2014;45:309–317. doi:10.1016/j.neuro.2014.05.006

12. Calderón-Garcidueñas L, Franco-Lira M, Henríquez-Roldán C, et al. Urban air pollution: Influences on olfactory function and pathology in exposed children and young adults. Experimental and Toxicologic Pathology. Published online 2010. doi:10.1016/j.etp.2009.02.117

13. Hudson R, Arriola A, Martínez-Gómez M, Distel H. Effect of Air Pollution on Olfactory Function in Residents of Mexico City. Chemical Senses. 2006;31(1):79–85. doi:10.1093/chemse/bjj019

14. Ajmani GS, Suh HH, Pinto JM. Effects of Ambient Air Pollution Exposure on Olfaction: A Review. Environmental health perspectives. 2016;124(11):1683–1693. doi:10.1289/EHP136

15. Cheng H, Saffari A, Sioutas C, Forman HJ, Morgan TE, Finch CE. Nanoscale particulate matter from urban traffic rapidly induces oxidative stress and inflammation in olfactory epithelium with concomitant effects on brain. Environmental Health Perspectives. Published online 2016. doi:10.1289/EHP134

16. Oleszkiewicz A, Pozzer A, Williams J, Hummel T. Ambient air pollution undermines chemosensory sensitivity – a global perspective. Sci Rep. 2024;14(1):30462. doi:10.1038/s41598-024-75067-z

17. Scussiatto HO, Da Silva JLB, Figueiredo AF, et al. Association of air pollution with olfactory identification performance of São Paulo residents: a cross-sectional study. Int Arch Occup Environ Health. 2023;96(4):621–628. doi:10.1007/s00420-023-01956-x

18. Hummel T, Power Guerra N, Gunder N, Hähner A, Menzel S. Olfactory Function and Olfactory Disorders. Laryngorhinootologie. 2023;102(S 01):S67-S92. doi:10.1055/a-1957-3267

19. Whitcroft KL, Cuevas M, Haehner A, Hummel T. Patterns of olfactory impairment reflect underlying disease etiology. The Laryngoscope. 2017;127(2):291–295. doi:10.1002/lary.26229

20. Wysocki CJ, Gilbert AN. National Geographic Smell Survey: Effects of Age Are Heterogenous. Ann NY Acad Sci. 1989;561(1 Nutrition and):12-28. doi:10.1111/j.1749-6632.1989.tb20966.x

21. U. S. EPA. National Ambient Air Quality Standards (NAAQS) \textbar Air and Radiation \textbar US EPA. *US Environmental Protection Agency*, Office of Air Quality Planning and Standards. Published online 2012.

22. Lee B. Highlights of the Clean Air Act Amendments off 1990. Journal of the Air & Waste Management Association. 1991;41(1):16–19. doi:10.1080/10473289.1991.10466820

23. Montreal Protocol on Substances that Deplete the Ozone Layer Final Act 1987. J Environ Law. 1989;1(1):128-136. doi:10.1093/jel/1.1.128

24. US Environmental Protection Agency Air Data. https://aqs.epa.gov/aqsweb/airdata/download_files.html

25. US Census Bereau. 2000 Census Zipcode Tabulation Areas. Published online 2000. https://www2.census.gov/geo/tiger/TIGER2010/ZCTA5/2000/https://www2.census.gov/geo/tiger/TIGER2010/ZCTA5/2000/

26. Bivand R, Rundel C, Pebesma E, Hufthammer KO. rgeos: Interface to Geometry Engine–Open Source (GEOS). R package version 01-8. Published online 2011.

27. Bivand R, Keitt T, Rowlingson B, Pebesma E. rgdal: Bindings for the geospatial data abstraction library. Published online 2019.

28. Pebesma E. Simple features for R: Standardized support for spatial vector data. R Journal. Published online 2018. doi:10.32614/rj-2018-009

29. Frye RE. Dose-Related Effects of Cigarette Smoking on Olfactory Function. JAMA. 1990;263(9):1233. doi:10.1001/jama.1990.03440090067028

30. Sorokowski P, Karwowski M, Misiak M, et al. Sex Differences in Human Olfaction: A Meta-Analysis. Front Psychol. 2019;10:242. doi:10.3389/fpsyg.2019.00242

31. Stata Press. Stata Statistical Software: Release 16. StataCorp LLC. Published online 2019.

32. Williams R. Generalized ordered logit/partial proportional odds models for ordinal dependent variables. Stata Journal. Published online 2006. doi:10.1177/1536867x0600600104

33. Benjamini Y, Hochberg Y. Controlling the False Discovery Rate: A Practical and Powerful Approach to Multiple Testing. Journal of the Royal Statistical Society: Series B (Methodological). Published online 1995. doi:10.1111/j.2517-6161.1995.tb02031.x

34. Knaapila A, Zhu G, Medland SE, et al. A Genome-Wide Study on the Perception of the Odorants Androstenone and Galaxolide. Chemical Senses. 2012;37(6):541–552. doi:10.1093/chemse/bjs008

35. Huang S, Li H, Wang M, et al. Long-term exposure to nitrogen dioxide and mortality: A systematic review and meta-analysis. Science of The Total Environment. 2021;776:145968. doi:10.1016/j.scitotenv.2021.145968

36. Jo S, Kim YJ, Park KW, et al. Association of NO_2_ and Other Air Pollution Exposures With the Risk of Parkinson Disease. JAMA Neurol. 2021;78(7):800. doi:10.1001/jamaneurol.2021.1335

37. Calderón-Garcidueñas L, Valencia-Salazar G, Rodríguez-Alcaraz A, et al. Ultrastructural Nasal Pathology in Children Chronically and Sequentially Exposed to Air Pollutants. Am J Respir Cell Mol Biol. 2001;24(2):132–138. doi:10.1165/ajrcmb.24.2.4157

38. Chew S, Lampinen R, Saveleva L, et al. Urban air particulate matter induces mitochondrial dysfunction in human olfactory mucosal cells. Part Fibre Toxicol. 2020;17(1):18. doi:10.1186/s12989-020-00352-4

39. Duan Y, Liao Y, Li H, et al. Effect of changes in season and temperature on cardiovascular mortality associated with nitrogen dioxide air pollution in Shenzhen, China. Science of The Total Environment. 2019;697:134051. doi:10.1016/j.scitotenv.2019.134051

40. Guan Y, Xiao Y, Chu C, Zhang N, Yu L. Trends and characteristics of ozone and nitrogen dioxide related health impacts in Chinese cities. Ecotoxicology and Environmental Safety. 2022;241:113808. doi:10.1016/j.ecoenv.2022.113808

41. Jhun I, Coull BA, Zanobetti A, Koutrakis P. The impact of nitrogen oxides concentration decreases on ozone trends in the USA. Air Qual Atmos Health. 2015;8(3):283–292. doi:10.1007/s11869-014-0279-2

42. Hasnain MG, Garcia-Esperon C, Tomari YK, et al. Effect of short-term exposure to air pollution on daily cardio- and cerebrovascular hospitalisations in areas with a low level of air pollution. Environ Sci Pollut Res. 2023;30(46):102438–102445. doi:10.1007/s11356-023-29544-z

43. Ajmani GS, Suh HH, Wroblewski KE, et al. Fine particulate matter exposure and olfactory dysfunction among urban-dwelling older US adults. Environmental Research. 2016;151:797–803. doi:10.1016/j.envres.2016.09.012

44. Zhang Z, Rowan NR, Pinto JM, et al. Exposure to Particulate Matter Air Pollution and Anosmia. JAMA Netw Open. 2021;4(5):e2111606. doi:10.1001/jamanetworkopen.2021.11606

45. Boogaard H, Patton AP, Atkinson RW, et al. Long-term exposure to traffic-related air pollution and selected health outcomes: A systematic review and meta-analysis. Environment International. 2022;164:107262. doi:10.1016/j.envint.2022.107262

46. Munger SD, Zhao K, Barlow L, et al. Towards Universal Chemosensory Testing: Needs, Barriers and Opportunities. Published online February 28, 2025. doi:10.31219/osf.io/5knb2_v1

47. Wise PM, Wysocki CJ, Lundstrom JN. Stimulus Selection for Intranasal Sensory Isolation: Eugenol Is an Irritant. Chemical Senses. 2012;37(6):509–514. doi:10.1093/chemse/bjs002

